# A thematic analysis of Prison and Probation Ombudsman fatal incident reports involving prisoners on palliative/end-of-life care pathways in the long-term high security estate in England and Wales

**DOI:** 10.1101/2021.06.21.21259231

**Authors:** Joanne Kirkham

## Abstract

**Objectives:** In this study, the clinical and non-clinical factors that may influence the provision of palliative/end of life care in long-term high security prisons in England and Wales are identified through the lens of Prison and Probation Ombudsman (PPO) fatal incident reports.

**Methods:** This work extends that of McParland and Johnston (2019) and contemporary literature published in the subsequent period to 2020 through a thematic analysis of fatal incident reports published by the PPO in the period 2014-2020. The results are discussed in context of the ‘Dying Well in Custody Charter’ and positioned within the extant literature.

**Conclusions:** The results suggest that prisoners in long-term high security prisons who are receiving palliative care are more likely than not to receive care that is comparable to that in the community. Directions for further research are also identified.

## Introduction

The prison environment is a complicated practice setting in the context of palliative/end-of-life care; clinicians must strive to deliver compassionate, consistent, and effective care within constraints that do not, ordinarily, exist in the community setting. Whilst the principle of equivalence of healthcare is now well embedded in and across the prison service in England and Wales, the ‘dying well in custody charter’ recognises the idiosyncrasies that challenge clinicians’ ability to respect the fundamental aspects of human dignity for prisoners who are receiving end-of-life nursing whilst serving a custodial sentence.

Older prisoners have, historically at least, accounted for a growing proportion of the prison population in England and Wales; 7% of prisoners were 50 years old or more in 2002, rising to 16% in 2018, thus representing the fastest growing age group (Sturge, G, 2018). Recent data suggests that the proportion has now stabilised at 16.6% (Ministry of Justice, 2020). Changes in sentencing conventions such as increased tariff and sentence length, Indeterminate Sentences for Public Protection (IPP) and prosecutions for historic sexual offences have contributed to the current population profile. As a result, the demand for healthcare services amongst the older population has increased, including for palliative and end-of-life care.

Evidence of the high level of physical and mental health problems experienced by older prisoners is summarised in ‘*Health and Social Care Needs Assessments of the Older Prison Population*’ (Public Health England, 2017); it recognises that that length of time in prison is a determinant of health (as well as age) and that those serving long-tariff sentences tending to experience poorer health outcomes than those of similar age who are incarcerated for much shorter periods. Hayes et. al. (2012) in PHE (2017) reports that up to 90% of the older prison population have one or more moderate to severe health conditions, and that > 50% present with three or more. The prevalence of chronic diseases is more evident amongst people in prison than in their ‘age-matched community peers and younger people in prison’ (PHE, 2017). This, coupled with a physical environment that is often not conducive to the long-term care of patients with chronic disease may place prison staff in the un-envious position of managing an ageing prison population and their associated age-related conditions in a clinical setting that may exacerbate, rather than moderate, the onset of further physical and mental health problems.

The consequence of an ageing prison population and changes in sentencing policy is an increased demand for palliative/end-of-life care. In recognition of this and the wider conversation around the role of hospices in the prison setting (see Markham, 2021), national guidance has sought to drive improvements in palliative/end-of-life care in prisons, and enable parity of care, commensurate with what may be reasonably expected by the general population in the community.

The Prisons and Probation Ombudsman (PPO), a non-statutory body in the UK that is responsible for, inter alia, investigations into deaths in custody has increasingly sought to cast a light on the provision of palliative/end-of-life care in prisons. It’s recent publication ‘*Learning from PPO investigations Older Prisoners*’ (2017), calls for a national strategy to address the needs of older people in prison. Amongst its recommendations of relevance to palliative and end-of-life-care, the report suggests that prisons should ensure that terminally ill prisoners who require intensive palliative care are treated in a suitable (physical) environment, in consultation with the prisoner, and that prisons should ensure that end of life and palliative care plans are commenced at an appropriate and early stage, recognising the vital need for advanced care planning.

In 2018, the ‘*Ambitions for Palliative Care and End of Life Framework*’ was adapted to reflect idiosyncrasies of the custodial setting. The resulting guidance, the ‘*Dying Well in Custody Charter*’, presents best practice recommendations and a set of guidelines relevant to each of the six recognised ambitions; these mirror those in the framework for the general population; i) each person is treated as an individual, ii) each person gets fair access to care, iii) maximising comfort and wellbeing, iv) care is coordinated, v) all staff are prepared to care and vi) each community is prepared to help. There is no published evidence on the efficacy of the charter; the associated self-assessment tool developed to support its implementation was tested in a pilot study at HM Prison Littlehey in 2018, where a multi-disciplinary group of a nurse, a governor and a palliative medicine consultant deployed the self-assessment tool to review current work.

### McParland and Johnston (2019)

In their mixed-methods, rapid review of the literature, McParland and Johnston (2019) seek to explore the current practices of palliative and end of life care in prisons and make recommendations for improvement in prisoner care. The review engages with the prisoner voice, their families, prison officers and prison healthcare staff as the basis for generating insights into;

- Current practices in palliative and end of life care.
- Barriers and facilitators to providing palliative and end of life care in prisons.
- The role of hospices.
- Recommendations for future provision of palliative and end of life care in prisons.

The paper is influenced by earlier work in Tricco et al (2015), where rapid reviews are advocated as a suitable form of synthesis; however, such approaches are, by nature limited and may lead to simplification at the expense of methodological rigour and therefore findings should be treated with caution. The review captures 23 published papers in total, which suggests that the extant literature is rather limited in this field.

McParland and Johnston’s review of the literature engages with current professional practice in prison healthcare settings. Themes that emerge include the very specific differences (and challenges) that are created by the nature of the prison environment, caring for the individual, including disregarding the background to the sentence, supporting, and managing prisoner volunteers and dealing with conflict of interests (e.g., security, discipline etc.) with prison officers and governors.

Papadopoulos and Lay’s 2016 online survey investigates the experiences of current and former prison nurses involved with the delivery of end-of-life care in UK prisons. Of the 31 respondents to the study, 68% stated that they had undertaken some training in end of life or palliative care, either due to prior clinical experience or short continuing professional development courses. The study characterises several barriers to effective end of life care: flexibility in prison regimes, lack of privacy, availability of appropriate care, visiting facilities and inadequate visiting hours. The relatively small sample size and anonymity of respondents, coupled with the problem of inconsistency in more than one respondent reporting on an individual prison, limits the generalisability of the study’s findings to the wider population of prisons and custodial settings in the UK.

McParland and Johnston’s review problematizes the challenges in providing palliative care in prisons through a bifurcation of barriers; physical and ideological. The prison environment itself is recognised as the most common physical barrier, examples include noisy landings or residential areas, problems with temperature regulation and humidity, staircases, and long corridors. Collaboration between the Marie Curie Hospice Edinburgh and HMP Edinburgh in 2018 recognises the prison environment and regime as an obstacle to providing effective palliative care in prisons. Their approach aims to reform palliative care for prisoners in HMP Edinburgh through a partnership approach with prison, health, and palliative care staff. The multi-disciplinary team committed to addressing some of these barriers by ensuring increased access to medication and subcutaneous infusion pumps when needed and exploring models to provide ‘Out of Hours’ and social care support. Other physical barriers in the literature review included drug administration and prescription and administration challenges. Two studies also found the prison regime a physical barrier, for example restricted visiting by family and friends. An Australian study (Panozzo et. al. 2020) explores the perceptions of health professionals providing palliative care for prisoners also found difficulties in pain and symptom management for end-of-life prisoners, with participants acknowledging the complex nature of palliative care in a prison environment.

Alongside the physical barriers to providing good palliative care in prisons the literature review examines ideological barriers, where two very different worlds collide, providing palliative care in the context of a custodial setting. The stark contrast between the two cultures culminates in examples of locked cell doors at end of life, the use of restraints and handcuffs during hospital appointments and public perceptions of the judicious use of public funds to improve a prisoner’s palliative care outcomes.

The Dying Well in Custody Charter (2018) places significant emphasis on each person being seen as an individual, including advanced care planning, ensuring the individual and the people important to them, are included in decision making. Three studies in McParland and Johnston’s literature review place importance on planning for end of life (Handtke et al, 2014, Sanders et al, 2018a and Sanders et al, 2018b). They recognise that prisoner’s daily lives are severely restricted and out of their control, which may increase the significance of advanced care planning.

The review suggests that prisoner relationships with families and other prisoners is a factor in promoting good palliative care. Examining the PPO reports from prisoners who received palliative care showed many viewed the prison as their home, expressing their preferred place of death as prison, despite other options such as compassionate release or hospice care. McParland and Johnston also found this to be true, with three studies classing their fellow inmates as a substitute family. A 2020 Belgian study by Humblet focused on the ‘emotional labour of prison professionals in the wake of an increasing number of older adults’. The study discussed how prison staff might struggle with the burden of care, particularly given the highly charged nature of a custodial setting. Humblet concludes that further research is required, to examine the emotional labour of prison staff towards prisoners who are, to some extent, viewed as a surrogate family.

Access to specialist services is a fundamental part of good palliative care. The ‘Dying Well in Custody Charter’ emphasises in the second ambition, that ‘each person gets fair access to care’. This ambition is designed to encourage greater awareness of how to access specialist support and advice. Part of ambition 6 of the charter, ‘each community is prepared to care’, presents guidelines that prisons should have agreements with local palliative care providers, including local specialist palliative care services and hospices. Both prisoners and clinicians in two studies from the literature review found this important. The survey by ==Popadopoulos and Lay (2016) also found access to specialist palliative care services as good practice in end-of-life care.

McParland and Johnston recognise the limitations of a rapid review within their report; the limited date range of 2014 – 2018 and a reduced number of databases searched. They acknowledge that some articles may have been missed from the review. New findings in their review included inmate hospice volunteers in delivering care to prisoners at end of life. These findings were not examined in this analysis to keep the focus UK based, as the studies involving this area were American based. The relevance of inmate volunteers is restricted, as not currently practice in UK prisons.

### Aim and objectives of the study

The aim of this study is to generate new evidence and identify opportunities for the improvement of palliative care provision in high-security prison settings in England and Wales. The aim is achieved through four objectives:

1. To critically review the literature on palliative care in the custodial setting in the period 2019-2021, through the lens of the ‘Dying Well in Custody Charter’.
2. To analyse PPO ‘Death in Custody’ investigation reports in cases where prisoners were receiving palliative care in the Long-Term High Security Estate in England and Wales.
3. To generate a thematic analysis using data from ‘Death in Custody’ investigation reports to identify clinical and non-clinical recommendations of relevance to Long-Term High Security Estate in England and Wales.
4. To locate the findings of the thematic analysis in the extant literature and present recommendations for i) improvement in the practice setting and ii) further research

### Research Methodology

A qualitative methodology is used in this study, comprising a short literature review building upon the existing literature described in McParland and Johnston (2019) and through analysis of PPO reports. The data collection and analysis is comprised of five stages: -

1. Identification of all PPO reports published from 2004 to 2020 in the long-term high security estate that are not self-inflicted, homicide or non-natural causes.
2. Keyword search in each report to identify the frequency of the term ‘palliative’ (this enabled the author to identify reports featuring sufficient information and data in relation to palliative care).
3. All reports outside the period 2014 to 2020 were removed from the analysis to match time frame of the literature review.
4. Tabulation of clinical and non-clinical concerns and recommendations from reports.
5. Results analysed against the 6 ambitions in ‘Dying Well in Custody Charter’ and the subsequent discussion section places the results of the thematic analysis in context of the ‘Dying well in Custody’ ambitions and the wider literature discussed in the review.

### Data collection

The Prisons and Probation Ombudsman (PPO) is a non-statutory body, part of the National Preventive Mechanism (NPM), and carries out independent investigations into complaints and deaths in custody in England and Wales. Amongst its core duties, the PPO investigates deaths of prisoners, young people in detention, approved premises’ residents, and immigration detainees due to any cause, including any apparent suicides and natural causes. The purpose of these investigations is to ‘understand what happened, to correct injustices and to identify learning for the organisations’ so that the PPO ‘makes a significant contribution to safer, fairer custody and offender supervision’.

In this study, all published PPO fatal incident reports at long-term high security prisons were accessed via the PPO website for the period 2004-2020. This represents the entire population of published reports. Due to the time frame of the literature review and following an examination of the reports, only those published between 2014 t0 2020 were taken forward for analysis. The clinical and non-clinical concerns were examined for each prison then assigned to the relevant theme in the ‘dying well in custody charter’ ambitions.

### Long-term high security prisons (LTHSE)

The Long Term and High Security Estate is comprised of 13 state run prisons housing the most dangerous and high-risk men and young people in the country. The LTHSE encompasses 8 High Security Prisons, one Young Offender Institution and four long-term ‘Category B’ prisons; HMYOI Aylesbury and HMP Belmarsh are located in London, HMP Frankland in County Durham, HMP Full Sutton in the East Riding of Yorkshire, HMP Garth near Preston, HMP Gartree near Market Harborough, HMP Manchester, HMP Isle of Wight (previously HMP’s Parkhurst, Albany and Camp Hill), HMP Swaleside on the Isle of Sheppey, HMP Wakefield, HMP Whitemoor in Cambridgeshire, HMP Long Lartin in Evesham, and HMP Woodhill in Milton Keynes.

## Discussion and limitations of the study

The findings from the analysis of the PPO reports reflect the many concerns identified in the surveyed literature, such as the importance of advanced care planning, access to specialist palliative care and communication between external services and the multi-disciplinary team. PPO reports included in this study highlight numerous examples of good practice in palliative care. From 71 reports examined, only 2 (2.8%) suggest that prisoners did not receive care equivalent to that in the community. It should be noted that the concept of equivalence of care is difficult to measure and is influenced by a range of factors that may be specific to the setting. In one report, the clinical reviewer ‘considered that healthcare staff provided the prisoner with coordinated and compassionate care’, which suggests that reviewers are mindful of the holistic need of patients. We summarise the evidence, drawn from the thematic analysis, through the prism of the 6 ‘Dying Well in Custody’ ambitions in Table 2.

**Table 1:**
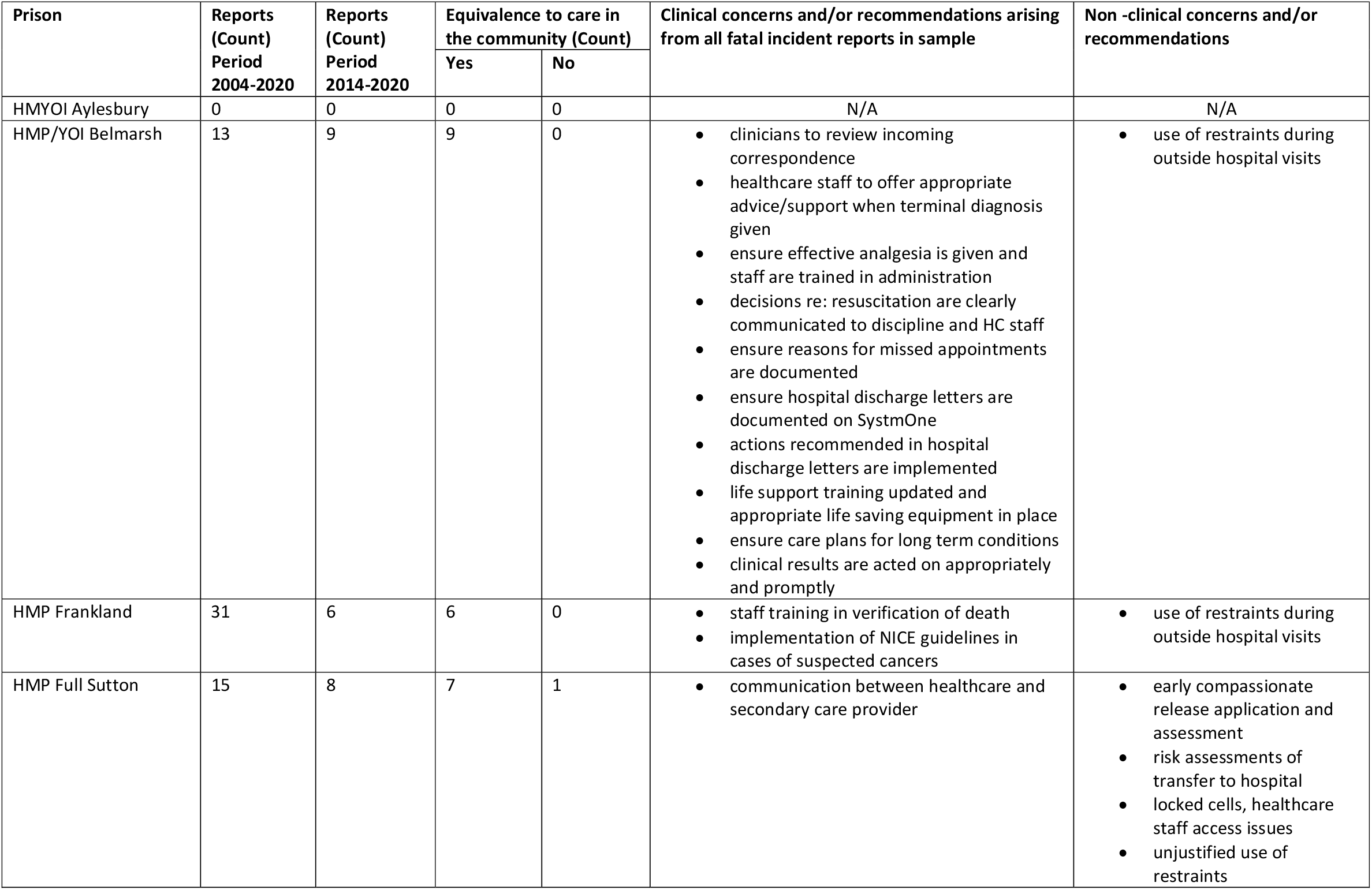
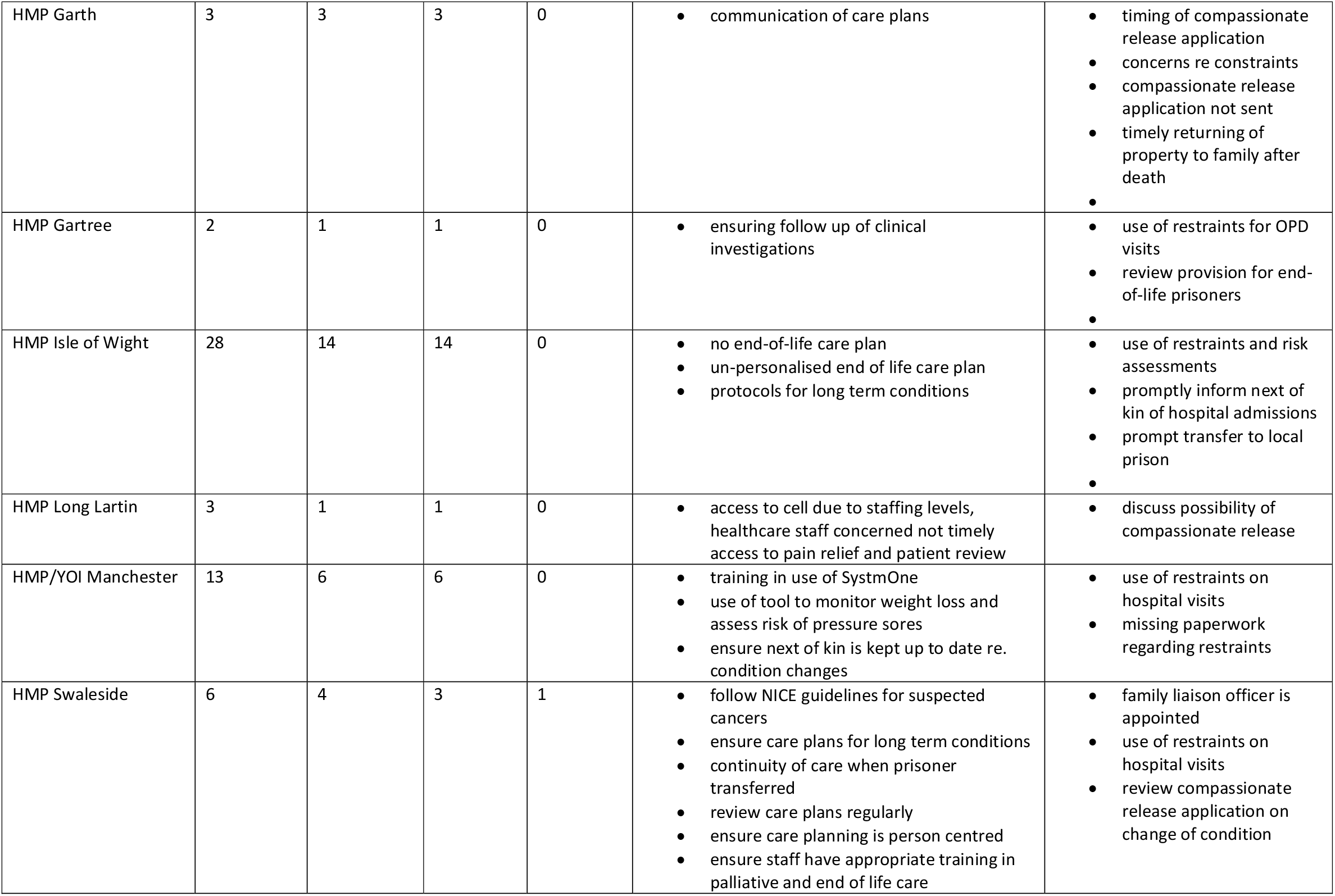
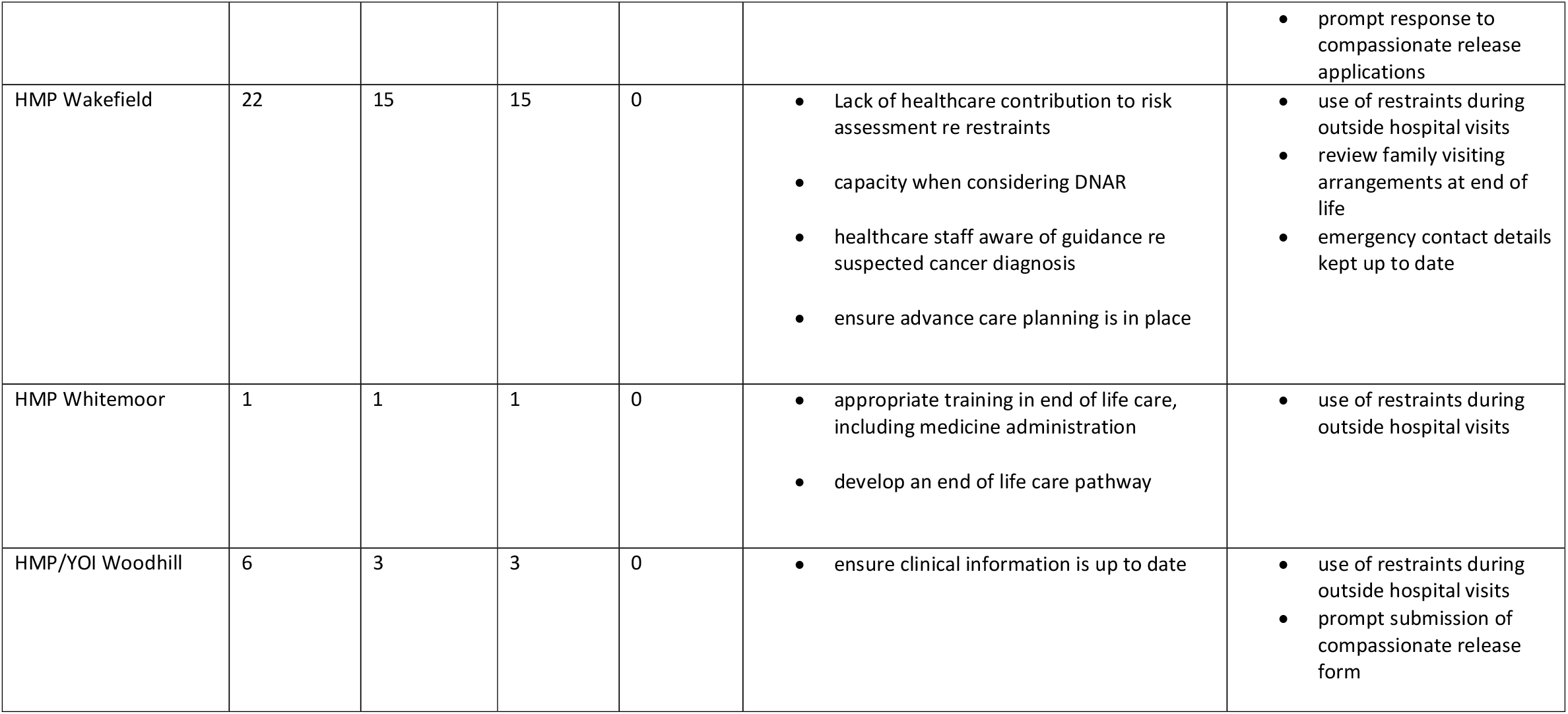
Results of thematic analysis of PPO fatal incident reports.

**Table 2:** Analysis of evidence arising from PPO investigation reports arranged by dying well in custody ambitions.

|  | **Dying well in custody ambitions** | **Evidence from PPO investigation reports** |
| --- | --- | --- |
| **1** | Each person is seen as an individual | Three reports identified deficiencies with advanced care and person-centred planning. Two reports identified deficiencies with DNAR decisions: one in relation to capacity and one in relation to information sharing. |
| **2** | Each person gets fair access to care | Two reports concluded that the person did not receive care that was considered equivalent to that what might reasonably be expected in the community setting. |
| **3** | Maximising comfort and wellbeing | In two cases concerns were identified in relation to drug administration. In one case, the use of tools to identify risk factors e.g. pressure sores. |
| **4** | Care is coordinated | Weaknesses in communication of care plans were identified in three cases. The appropriate documentation of information on SystemOne was identified in two cases. |
| **5** | All staff are prepared to care | There was no evidence in the reports of concerns in relation to this ambition. The reports contain numerous examples of excellent care, and in some cases, above what may have been provided in the community. |
| **6** | Each community is prepared to help | Good links to local hospices are identified in several reports. Recommendation for Ministry of Justice to consider the estate and its fitness for purpose for palliative prisoners and ageing prison population profile. |

The limitations of this study are grounded in the use of a single source of data (PPO reports) and the assumption that the PPO is competent to assess if the treatment of patients in a custodial setting is comparable to the care that they might reasonably expect to receive in the community. In respect of the former, further research should seek to gather and triangulate evidence from Independent Monitoring Board (IMB) annual reports, HM Inspector of Prisons (HMIP) reports and Care Quality Commission (CQC) reports.

## Conclusion

‘Learning from PPO Investigations Older Prisoners’ (2017), identifies two lessons learned in palliative and end of life care, arising in PPO fatal incident investigations into the death of older prisoner from 2013 to 2015. Firstly, they identify that ‘prisoners who need intensive palliative care are treated in a suitable environment, in consultation with the prisoner’, and secondly, the lesson learnt is relating to advanced care planning and care plans should ‘include all aspects of patient care, including effective pain relief and psychological and emotional support’. The suitable environment is difficult to establish from the PPO reports analysed, although with only two incidents of care not comparable to community based, it appears efforts are being made to facilitate the patient’s needs. The evidence from this analysis confirmed advanced care planning remains an area of concern within a palliative care context in prison. National Institute for Care and Excellence guidelines for end-of-life care for adults (2011) ensure ‘people approaching the end of life are offered comprehensive holistic assessments in response to their changing needs and preferences, with the opportunity to discuss, develop and review a personalised care plan for current and future support and treatment’. Personalised advance care planning would assist in communication and coordination issues in providing good quality palliative care in prisons. Overall, it appears palliative care in high security prisons is comparable to community care, with links being fostered with specialist palliative care teams and hospices, efforts are being made to improve the care of this ageing population.

## Limitations of the study

PPO reports are framed through one lens of enquiry and may not always sufficiently capture the nuances of individual cases. There are other sources of data and evidence which may improve the granularity of the results including reports from the Care Quality Commission, Her Majesty’s Inspectorate of Prisons, and the Independent Monitoring Boards. Further research should seek to triangulate additional data sources to confirm the relevance and appropriateness of PPO findings.

## Data Availability

The study uses publicly accessible data

